# Measuring Youth Mental Health Vulnerability Across Communities: Development and Validation of ThriveAtlas

**DOI:** 10.64898/2026.01.26.26344733

**Authors:** Christina A. Mehranbod, Domonique M. Reed, Emily Yetton, Aaron Dibner-Dunlap, Tomas Engelthaler, Peter Smittenaar, Hannah Kemp, Sema Sgaier, Rui Adele H Wang

## Abstract

**Objective:** Youth mental health is shaped by interacting individual, relational, and structural factors that vary across communities, yet few tools integrate these drivers into youth-specific, geographically granular measures for decision-making. This study describes the development and preliminary validation of ThriveAtlas^TM^, a geographically granular index designed to identify factors that prevent youth from thriving.

**Methods:** ThriveAtlas was developed through a multi-step process to identify, operationalize, and validate contextual determinants of youth mental health and well-being. This process included a literature review; selection and construction of indicators from public and proprietary data; aggregation into thematic domains and a composite index; and preliminary validation using county-level mental health outcomes in two U.S. states. Principal components analyses using California and Washington data were used to derive dimensions of youth mental health, which were correlated with ThriveAtlas index and subthemes.

**Results:** The review identified key gaps in existing tools: youth-focused indices often emphasize general well-being rather than mental health, while structurally-focused indices lack youth specificity or sub-state granularity. ThriveAtlas revealed geographic variability in vulnerability, including regional clusters and isolated high-risk counties. Validation analyses showed expected patterns in California, where higher vulnerability aligned with greater distress and lower well-being. In Washington, correlations varied by outcome, with stronger correlations observed for crisis-related mental health indicators than overall distress.

**Conclusion:** ThriveAtlas addresses a critical measurement gap by providing a youth-centered, multidimensional, and geographically granular index of vulnerability. Early validation supports its utility as a decision-relevant signal to identify communities with elevated youth mental health challenges and inform targeted intervention strategies.

## Introduction

Adolescence and emerging adulthood are critical stages of development, marked by profound biological, psychological, and social changes that impact mental health.^1^ Most mental health conditions emerge during this period and often follow a chronic or relapsing course, with lasting effects on opportunities for a flourishing life.^2^ Evidence suggests that adolescents and young adults are experiencing a more severe mental health crisis compared to previous generations.^3^ Recent surveillance data indicate that mental health conditions affect approximately 20% of youth in the United States (U.S.) annually, with even higher proportions reporting mental health symptoms and emotional difficulties.^4–6^ This widespread crisis has far-reaching implications, including increasing burden on healthcare systems, straining school-based mental health services, and heightening the need for comprehensive familial, social, and community support programs.^7,8^

A substantial body of evidence demonstrates that youth mental health and well-being are shaped by interacting risk factors, including individual experiences, social and familial relationships, and structural conditions.^9–15,15–18^ These determinants are unevenly distributed across space, with cumulative risk often clustering geographically in ways that shape youth’s access to resources, exposure to stressors, and opportunities for support.^19,20^ As such, measurement strategies that integrate multiple dimensions of vulnerability into interpretable, geographic metrics are needed to support prioritization, resource allocation, and systematic monitoring over time.^21,22^ However, most existing indices were not designed specifically to capture youth mental health risk and vary widely in their inclusion of youth-relevant domains, systems (e.g., schools and youth-serving providers), and geographic scale.

Understanding what existing tools measure, and what they omit, is critical for advancing measurement innovation and developing frameworks that better capture the multidimensional nature of youth mental health.^23,24^ A range of tools has been developed to monitor burden and trends in mental health well-being, but these tools vary substantially in target audience, scope, and geographic coverage.^23,24^ National surveillance systems and youth-specific surveys provide valuable insight into population-level trends in mental health outcomes and related behaviors, offering epidemiologic insight into the scope of the crisis.^25,26^ Additional efforts, such as the social vulnerability index or the neighborhood disadvantage index incorporate key social and neighborhood determinants of health, while existing child well-being indices largely focus on physical, educational, or socioeconomic outcomes and give limited attention to youth mental and emotional health.^27–32^ However, across these tools, coverage of domains salient to youth mental health (e.g., peer and school climate; youth-specific mental health practitioners and providers) varies substantially, as does the geographic scale at which they operate. Collectively, these efforts have produced a fragmented landscape of measurement tools for youth mental health and well-being, with substantial variation in geographic resolution and domain coverage. This fragmentation reflects both the complexity of youth mental health as a construct and the difficulty of developing tools that are comprehensive, contextually sensitive, and actionable across diverse geographic settings.

This study describes the conceptual foundation, development process, and preliminary validation of ThriveAtlas^TM,^ a geographically granular index designed to identify factors that prevent youth from thriving.

## Methods

### Overview of ThriveAtlas Development

ThriveAtlas was developed through a multi-step process designed to identify, operationalize, and validate geographic-specific determinants of youth mental health and well-being. The development process consisted of: 1) a targeted literature review to identify relevant domains and existing tools measuring youth mental health and well-being; 2) selection and construction of indicators using publicly available and proprietary data sources; 3) aggregation of indicators into thematic domains and a composite index; and 4) preliminary validation of the index against adolescent mental health outcomes in two U.S. states. ThriveAtlas is a registered trademark of Surgo Health.

### Target Literature Review

#### Search strategy and database selection

We conducted a literature search between January 2010 to September 2025 across PubMed, PsychINFO, and Google Search. The search strategy combined terms related to youth and adolescents (e.g., “adolescent,” “young adult,” “youth”, “children”), mental health (e.g., “mental health”, “well-being”, “emotional”), geographic organization (e.g., “neighborhood,” “community,” “social environment”), determinants (e.g., “poverty,” “education,” “housing,” “school climate,” “family relationships,” “peer support,” “adverse childhood experiences,” “healthcare access”), and measurement tools (e.g., “index,” “composite indicator,” “score,” “tool”). To supplement database searches, gray literature, such as websites maintained by national health organizations and academic institutions were reviewed to identify reports, summary estimates, and proprietary tools not indexed in academic databases. References of included tools were examined to identify additional eligible material.

#### Eligibility Criteria

Tools were eligible for inclusion if they captured aggregated burdens or determinants of youth mental health and well-being, and focused on U.S. populations, and its sub-national geographic units, such as state, county, or ZIP code. Tools were excluded if they focused exclusively on individual-level risk factors or did not provide information on included domains, indicators, or data sources. These criteria aligned with ThriveAtlas’s emphasis on modifiable, geographically concentrated determinants relevant to policy and programmatic decision-making.

#### Extraction

For each eligible tool or framework, we extracted descriptive information including name, purpose, geographic scope, and domains. Extracted data were organized into summary tables to enable comparison and synthesis across the available tools (Table 1). A narrative synthesis was used to characterize the scope of included tools and their limitations. Insights derived from this synthesis were subsequently used to guide the development of the ThriveAtlas index.

**Table 1.** Selected Existing Geographically Granular Tools to Characterize Burden of and Risk Factors Associated with Mental Health and Well-being among Youth.

| Name<br>Type of Tool<br>Geographical Unit | Indicators | Criteria |  |  |  |
| --- | --- | --- | --- | --- | --- |
|  |  | Youth-focused | Mental health<br>*and* well-being<br>focused | Geographically<br>granular | Incorporate<br>underlying<br>drivers of<br>vulnerability for<br>actionability |
| Summary estimates describing burden of youth mental health and associated risk factors |  |  |  |  |  |
| Youth Risk Behavior Surveillance System<br><sup>46</sup><br>**Summary Estimates**<br>National, state, selected districts | Sadness or hopelessness; poor mental health; suicidal ideation and attempts; substance use; experience of violence, including physical and cyberbullying; parental monitoring; school connectedness; unstable housing; racism; and social media use | ✓ | ✓ | ✓ | X |
| National Survey of Children's Health<br><sup>49</sup><br>**Summary Estimates**<br>National, state, selected sub-state regions | Diagnosed mental health conditions; social and emotional well-being; health-related behaviors (e.g., sleep); health care access and use; school environment and engagement; adverse childhood experiences; and community activities and experiences | ✓ | ✓ | ✓ | X |
| National Survey | Substance use; symptoms of | ✓ | ✓ | ✓ | X |
| <u>on Drug Use and Health</u><br>48<br>**Summary Estimates**<br>National, state, sub-state | generalized anxiety disorder; major depressive episode or any mental illness; suicidal ideation and attempts; substance use treatment; mental health treatment |  |  |  |  |
| <u>National Survey on the Mental Health of LGBTQ+ Young People</u><br>47<br>**Summary Estimates**<br>State | Access to mental health care; violence and bullying; discrimination; political climate; family support; peer support; suicide risk; anxiety and depression | ✓ | ✓ | X | X |
| <u>County Health Rankings and Roadmaps</u><br>51<br>**Summary Estimates**<br>County | Children in poverty; uninsured children; children eligible for free or reduced lunch prices; children in single-parent household; mental health providers | ✓ | ✓ | ✓ | X |
| <u>America's Health Rankings</u><br>50<br>**Summary Estimates**<br>State | Children's mental health treatment; household smoke around child; child victimization; children in poverty; students experiencing homelessness; education; youth soda consumption; children adequately insured; tobacco, vape, or illicit drug use in children; alcohol use in children; adverse childhood experiences; foster care instability; neighborhood amenities; residential segregation; adequate sleep; food sufficiency; high-speed internet; physical activity; obesity; teensuicide | ✓ | ✓ | X | ✓ |
| **Index to identify geographic regions where youth mental health is not thriving** |  |  |  |  |  |
| <u>State of Mental Health in America Ranking (Youth)</u><br>52<br>**Ranking tool**<br>State | Major depressive episode; substance use disorder; suicidal ideation; flourishing; inadequate private insurance coverage for mental and emotional problems; health care access; individualized education programs | ✓ | ✓ | X | ✓ |
| **Index to identify geographic regions where youth well-being is not thriving (lacks emphasis on mental health)** |  |  |  |  |  |
| <u>Child and Youth Well-Being Index</u><br>31<br>**Index**<br>National, State<sup>a</sup> | Family economic well-being, health, and social relationships; social relationships; general health; health and safety/risk behaviors; educational attainment; emotional/ spiritual well-being | ✓ | X | X | ✓ |
| <u>KIDS COUNT Child Well-being in the United States</u><br>32<br>**Index** | Economic well-being; general health and insurance coverage; education; and family and community | ✓ | X | X | ✓ |
| State |  |  |  |  |  |
| <u>Child Opportunity Index</u><br>29<br>**Index**<br>National, state, metro, county, zip code, census tract | Education and educational resources; pollution, healthy environments, safety-related resources, health resources; employment, economic resources, socioeconomic inequity, housing resources, social resources, and wealth | ✓ | X | ✓ | ✓ |
| <u>Child and Adolescent Thriving Index 1.0</u><br>27<br>**Index**<br>National, state | Non-low birth weight; preschool attendance; reading proficiency; math proficiency; food security; general health status; non-obesity; non-smoking; non-marijuana use; non-arrest; high school graduation | ✓ | X | X | ✓ |
| **Index to identify neighborhoods and communities that are structurally vulnerable** |  |  |  |  |  |
| <u>Social Vulnerability Index</u><br>(CDC, 2024)<br>**Index**<br>County, zip code | Socioeconomic status; household characteristics; racial and ethnic minority status; and housing and transportation | X | X | ✓ | X |
| <u>Area Deprivation Index</u><br>(Kind & Buckingham, 2018)<br>**Index** | Income; education; employment; and housing quality/conditions | X | X | ✓ | X |
| County, zip code,<br>census block |  |  |  |  |  |
| <sup>a</sup>State-specific indices have been developed based on this index (e.g., California Index of Child and Youth Well-being); |  |  |  |  |  |

### Indicator Selection and Construction

#### Indicator Selection Criteria

To guide indicator selection for ThriveAtlas, we applied five criteria: importance, actionability, data availability, stakeholder relevance, and equity, to identify factors from the literature for inclusion in the index, ensuring scientific rigor and policy relevance. Importance reflects both prevalence of a factor and the strength of its association with mental health outcomes. Actionability ensures that included factors are modifiable and offer potential intervention points for policymakers and practitioners. Data availability assesses whether a factor can be measured reliably and consistently across geographies or populations using existing data sources. When geographically granular data were limited, spatial estimation methods were applied to generate stable local estimates (Supplemental Material S1).^33^ Stakeholder relevance prioritizes factors aligned with the informational needs of policymakers, community leaders, and youth-serving organizations. Equity ensures that factors salient for marginalized youth are represented, preventing the framework from centering more privileged populations and highlighting structural inequities relevant to youth mental health.

#### Data Sources

Using these criteria, we identified indicators from publicly available data sources such as the American Community Survey, Behavioral Risk Factor Surveillance System, Civil Rights Data Collection, as well as Surgo Health’s Provider Database (Supplemental Material S2). When multiple sources were available, we prioritized those that were most complete, recent, and geographically granular. Indicator selection incorporated both compositional variables, which reflect population characteristics aggregated from individual-level data, and contextual variables, which capture place-based structural or environmental conditions shaping exposures and opportunities, independent of individual attributes. Indicators were available for 97.4% of counties included in the analysis.

#### Indicators Weighting

Assigning differential weights to indicators is challenging and potentially subjective, as there is no definitive or comprehensive evidence base (e.g., meta-analysis) that estimates the relative population-level impact of all indicators included in the index.^34^ Implementing differential weights would therefore require strong assumptions about the relative importance of individual drivers.^35^ For example, adverse childhood experiences are well established as strong predictors of mental health outcomes at the individual level, yet they occur at substantially lower prevalence than other drivers, like lack of emotional support, complicating their translation into population-level, place-based weights. Moreover, youth mental health risk does not arise from simple one-to-one relationships between individual indicators and outcomes. The drivers included in the index are interrelated and often operate through bidirectional and reinforcing pathways, making isolated weighting of individual factors conceptually misleading.^14,36^

Given these considerations, we apply equal weighting as a conservative and transparent approach.^35–37^ However, a known limitation of equal weighting is that adding indicators within a subtheme or theme can dilute the influence of any single indicator.^34^ This consideration is explicitly incorporated into the index design process. Indicators are included only when there is robust evidence supporting their relevance to youth mental health vulnerability, and subthemes are intentionally constructed to avoid overloading, ensuring that each indicator remains meaningfully represented. Under this framework, the composite index is designed to capture the concentration and co-occurrence of multiple, well-established drivers of youth mental health vulnerability within a place, while minimizing subjective assumptions about relative importance.

#### Index Construction

Operationally, ThriveAtlas assigns each geography a composite score ranging from 0 to 100, where 0 represents the lowest and 100 the highest relative risk level. To balance analytical rigor with interpretability, ThriveAtlas combines a composite index structure with a relative ranking approach. Scores are generated using an iterative percentile-ranking procedure similar to those used in established indices.^38^ Each indicator value is first multiplied by its valence, where a valence of 1 denotes an indicator associated with greater risk (i.e., lower well-being) and −1 denotes an indicator associated with greater well-being, ensuring that higher values consistently reflect higher risk. Valence-adjusted indicator values are then percentile-ranked within each geographic unit. Indicator-level percentile scores are averaged within sub-themes and re-ranked, with all indicators weighted equally. This process is repeated at successive levels of aggregation (themes and the overall index), producing a final ThriveAtlas score for each geography on a 0–100 scale.We displayed the geographic distribution of the ThriveAtlas Index to examine variation by county in vulnerability across the U.S..

#### Index Validation

To validate the ThriveAtlas Index, we conducted a county-level analysis in California and Washington states to assess the correlation between geographic vulnerability and adolescent mental health outcomes. We chose these two states as they had publicly accessible data that were sufficiently complete, consistently defined, and temporally aligned with the index construction. We used data from the California Healthy Kids Survey (2021–2023) and the Washington State Healthy Youth Survey (2023), two statewide, school-based surveys administered biennially.^39,40^ For the California analysis, we examined responses from ninth-grade students across 53 of the state’s 58 counties, assessing five self-reported mental health indicators: suicidal ideation, social-emotional distress, optimism, life satisfaction, and depressive feelings. For the Washington analysis, we analyzed responses from tenth-grade students across 32 of the state’s 39 counties, assessing eight self-reported indicators: uncontrolled worrying; suicidal ideation, planning, and attempts; lack of trusted adults when feeling depressed; feeling nervous, anxious, or on edge; depressive feelings; and generalized anxiety disorder. Given the multidimensional nature of youth mental health, we assessed correlations among indicators within each state using the Spearman’s correlation coefficients (ρ).^41,42^ We then conducted state-specific principal components analysis (PCA) to reduce correlated indicators into a smaller set of components representing underlying dimensions of adolescent mental health.^43^ Prior to PCA, all outcomes were standardized using z-score transformations to account for differences in scale and variance, and to prevent any single indicator from disproportionately influencing component structure. Finally, Spearman’s ρ was used to assess correlation between ThriveAtlas index, subthemes, and the state-specific mental health outcomes, with coefficients interpreted as weak (±0.10–0.29), moderate (±0.30–0.49), or strong ( ±0.50–1.00).^44^

## Ethics approval

The authors assert that all procedures contributing to this work comply with the ethical standards of the relevant national and institutional committees on human experimentation and with the Declaration of Helsinki of 1975, as revised in 2008. This study did not involve primary data collection or any interactions with human subjects. All data sources were publicly available (e.g., American Community Survey) or not identifiable (e.g., provider database), and therefore did not require IRB review.

## Data Sharing

Data are available upon reasonable request. Limited data sharing may be considered. Requests should be sent to Surgo Health at.

## Results

### Literature Review

To support actionable decision-making, an adequate tool for assessing vulnerability to youth mental health should meet four core criteria: it should be explicitly youth-focused, center mental health and well-being rather than general disadvantage or broad health indicators, operate at a level of geographic granularity sufficient to capture and identify meaningful variation across communities, with county-level data as a minimum standard, and finer resolution where feasible. In addition, the tool should emphasize underlying drivers of vulnerability, not only the prevalence or severity of mental health challenges, so that decision-makers can move beyond identifying and describing where needs are greatest to understanding what factors can be addressed through policy and programmatic action.

Much of the existing literature examines single risk factors in isolation, such as neighborhood deprivation, poverty, or access to care, rather than the constellation of interacting influences that jointly contribute to mental well-being.^11–13,45^ Nationally representative surveys (e.g., Youth Risk Behavior Surveillance System, National Survey of Children’s Health), have been instrumental in producing aggregated prevalence estimates of mental health burden among youth, as well as many of the relevant risk and protective factors driving youth mental health (Table 1).^46–49^ Similarly, summary statistics published by governmental and advocacy organizations, provide valuable snapshots of state-level or county-level trends in mental health burden.^50,51^ While necessary, burden-focused tools alone offer limited guidance for action, as they do not identify the underlying factors that shape variation in youth mental health across communities.

Although broader public health indices, such as the Social Vulnerability Index (SVI) and NeighborhoodAtlas^Ⓡ^’s Area Deprivation Index (ADI), help identify communities facing structural disadvantage, they are not designed to capture the determinants specific to youth mental health.^28,30^ The SVI was designed to assess community vulnerability during public health emergencies, and the ADI captures neighborhood socioeconomic deprivation; both rely on broad structural indicators (e.g., socioeconomic status, household composition, housing conditions) and overlook youth-specific factors such as safety, belonging, and peer dynamics.^14^ As a result, using such indices may obscure important pathways linking geographic context to youth mental health.

Few existing tools are designed to characterize youth mental health and well-being in ways that reflect the unique developmental contexts of youth. When evaluated against criteria of youth focus, mental health specificity, and geographic granularity, most available tools reveal substantial gaps. We identified only one publicly available tool that captured youth mental health at an aggregated geographic level: The State of Mental Health in America Youth Ranking. This tool integrates measures of prevalence (e.g., rates of major depressive episodes, substance use disorder, and suicidal ideation among youth) with indicators of access to care, such as the proportion of youth receiving mental health services or having insurance that fails to cover mental or emotional health problems.^52^ This ranking approach identifies state-level disparities in mental health burden and access, offering a policy-relevant tool for prioritization; however, its reliance on a limited set of indicators captures a narrow dimension of youth mental health and lacks the granularity needed to detect within-state variation.

Several established youth-focused indices assess general well-being but emphasize physical, educational, or socioeconomic indicators rather than mental and emotional health.^27,29,31,32^ For example, the Child and Youth Well-Being Index aggregates indicators across economic, health, education, and community domains over time, with emotional and spiritual well-being comprising a relatively small component of the overall composite.^31^ Similarly, KIDS COUNT Child Well-being Index captures outcomes across four domains: economic well-being, education, health, and family and community context, but is limited to state-level analysis, a notable limitation given that risks and resources vary substantially across communities.^32^ The Child and Adolescent Thriving Index V0.1 centers positive developmental outcomes using indicators of physical health, academic achievement, and behavioral risk at national and state levels but largely reflects general functioning and risk avoidance, with limited attention to mental and emotional health or within-state variation.^27^ In contrast, the Child Opportunity Index provides fine-grained geographic estimates (e.g., national, state, metropolitan, county, ZIP code, and census tract-level) across three broad domains: education and education resources; health and environment; and social and economic opportunity.^29^ However, despite meeting criteria for geographic specificity, the index omits mental health-specific dimensions, including peer belonging, family functioning, interpersonal support, and other barriers that shape youths’ ability to access mental health care, limiting its relevance for youth mental health vulnerability.

Although existing tools reflect advances in child welfare measurement and support state-level comparisons, none are simultaneously youth-focused, mental health-centered, and geographically granular. This gap underscores the need for tools that integrate multiple developmental and contextual dimensions at fine geographic scales.

Given the limitations of the scope of these tools, we also reviewed literature to determine relevant factors to include, we conducted a literature review to identify factors beyond individual vulnerabilities that impact youth mental health. We identified the following factors as consistently associated with youth mental health in the literature: family relationships and dynamics, socioeconomic status, adverse childhood and life experiences, peer relationships and social support, physical health and wellness behaviors, community and neighborhood environment, school environment, and genetic and biological factors.

### ThriveAtlas Index

The ThriveAtlas addresses gaps identified in the literature by providing a relative vulnerability index across U.S. census tract, ZIP code, county, and state levels, including Washington, D.C., Alaska, and Hawaii. The index is constructed from 25 indicators grouped into six thematic domains relevant to youth mental health and well-being. These domains were defined based on empirical evidence, refined with input from subject matter experts, and contribute equally to the composite ThriveAtlas score (Table 2).

**Table 2.** ThriveAtlas Themes Description and Indicators.

| Themes and Subthemes |  | Indicators |
| --- | --- | --- |
| **Theme 1** | **Limited Wellness Practice**<br>Focuses on the behaviors that impact physical and mental wellness | Sleep |
|  |  | Exercise |
|  |  | Daily functioning |
| **Theme 2** | **Provider Shortage**<br>Addresses the capacity of a community's mental health infrastructure by examining the availability and variety of mental health providers and organizations. | Mental health organizations |
|  |  | Youth psychologists |
|  |  | Youth psychiatrists |
|  |  | Youth non-clinical mental health providers |
|  |  | School psychologists |
| **Theme 3** | **Accessibility Barriers**<br>Addresses the barriers that youth encounter when seeking mental healthcare | Households without internet |
|  |  | Transportation costs |
|  |  | Uninsured minors |
|  |  | Unaffordable medical visits |
|  |  | Youth mental health insurance coverage |
| **Theme 4** | **Socioeconomic Hardships**<br>Explores the socioeconomic factors, such as poverty, education, and employment, that influence mental health outcomes and access to mental healthcare. | Households below the poverty line |
|  |  | Education |
|  |  | Unemployment |
|  |  | Disconnected Youth |
| **Theme 5** | **Negative life experiences**<br>Addresses experiences that impact mental health | Family structure |
|  |  | Adverse childhood experiences |
|  |  | Neighborhood safety |
| **Theme6** | **Limited support and belonging**<br>Explores the quality and nature of relationships within families and peer groups, and social support networks | Emotional support |
|  |  | Social isolation |
|  |  | School staff |
|  |  | Bullying and harassment |
|  |  | Childcare facilities |

#### National Distribution of Vulnerability Score and Themes

Figure 1 maps county-level ThriveAtlas vulnerability scores across the U.S., revealing pronounced geographic inequities. The highest vulnerability is concentrated in the South, lower Midwest, and parts of the West. These patterns reflect clusters of structural and social disadvantage, indicating regions where barriers to youth thriving are most deeply entrenched. Although vulnerability is regionally concentrated, isolated high-risk counties appear nationwide, suggesting that elevated need is not confined to any single part of the country.

**Figure 1.**
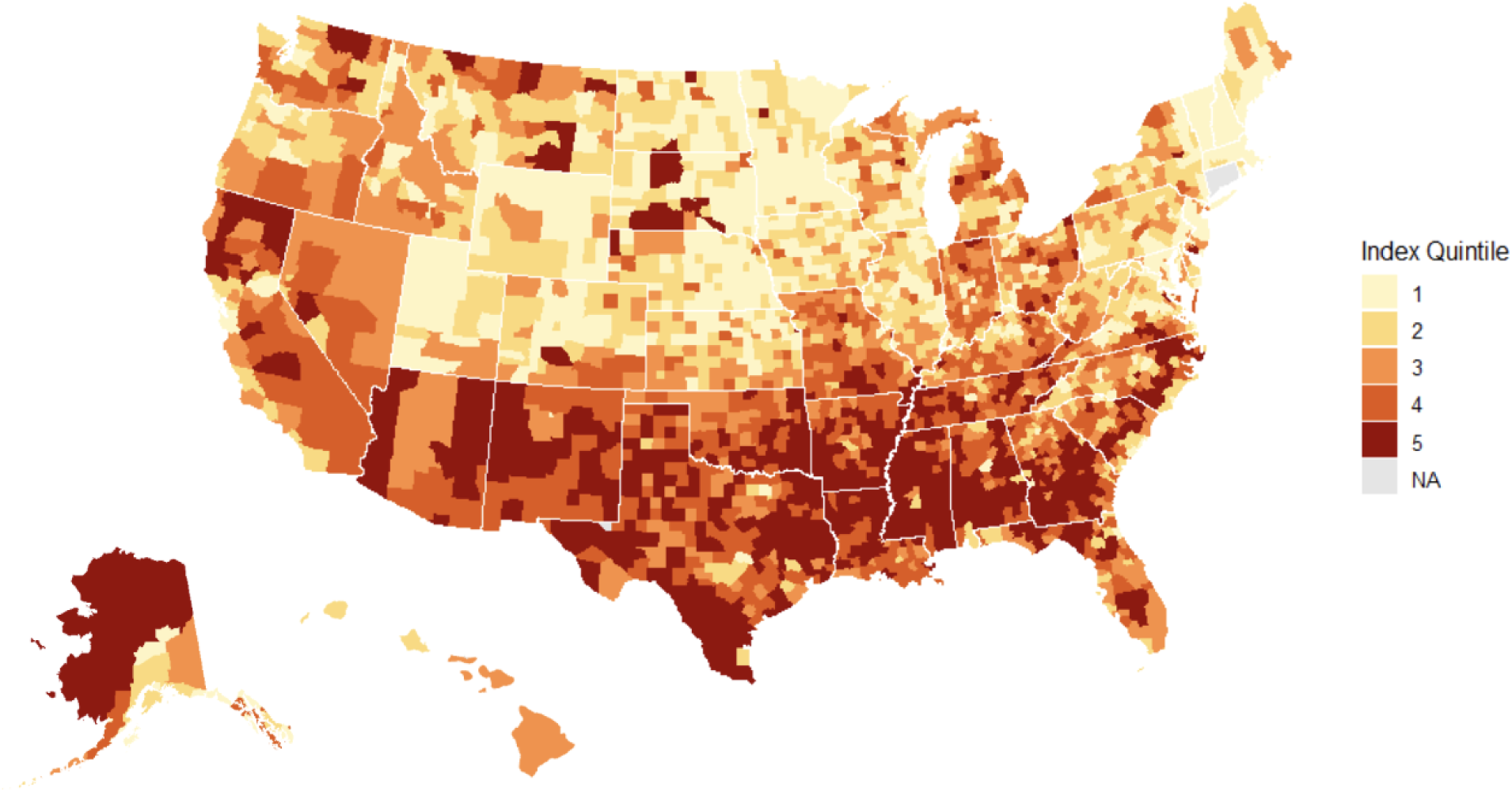
Map of the County-level Vulnerability Scores across the United States. Higher index scores indicate greater vulnerability.

#### State Validation

Across both states, negative mental health indicators were moderately to strongly correlated, while protective well-being indicators moved in the opposite direction (Supplemental Material S3 Figures S1 and Figure S3). In California, depressive feelings, social emotional distress, and suicidal ideation were strongly positively correlated (ρ ≥0.81; all p-value <0.05), whereas optimism and life satisfaction were moderately to strongly negatively correlated with these indicators (ρ ranged from −0.60 to −0.39; all p-value <0.05). In Washington, all of the outcomes were negative mental health indicators, and of those, internalizing symptoms, such as uncontrolled worry, anxiety, nervousness, and depressive feelings, were strongly correlated (ρ ≥0.70; all p-value <0.05), while indicators related to support such as having no adults to turn to when feeling depressed, showed weaker correlations (ρ ranged from 0.26 to 0.36), with mixed statistical support.

In California, PCA resulted in two dominant components jointly explaining 86.9% of the variance (Supplemental Material Figure S2 and Table S1). Component 1 represented a broad distress continuum, driven by high levels of depressive feelings, social-emotional distress, and suicidal ideation, contrasted with lower levels of life satisfaction and optimism. Component 2 captured a targeted well-being dimension, distinguishing counties characterized by particularly strong optimism and life satisfaction, independent of overall distress. In Washington, Component 1 explained 57.7% of the total variance, reflecting a unified internalizing distress factor, with anxiety, worry, depressive symptoms, and suicidal ideation all loading strongly (Supplemental Material Figure S4 and Table S2). Component 2 accounted for an additional 15.2% of the variance and differentiated acute crisis from anxiety, with suicide attempts and lack of adult support when feeling depressed defining one end of the component, and emotional distress symptoms on the other end, including feeling nervous or on edge, uncontrolled worrying, and generalized anxiety.

We correlated PCA components with the ThriveAtlas Index and subthemes (Figures 2a-b). In California, overall distress (Component 1) had a weak to strong positive correlation (ρ ranged from 0.18 to 0.64) with vulnerability themes, suggesting that higher county vulnerability is generally correlated with greater reported distress among 9th-grade students. The strongest correlations were observed for Accessibility Barriers (ρ = 0.64), Overall Index (ρ = 0.49), and Provider Shortages (ρ = 0.45). Overall well-being (Component 2) showed uniformly negative correlations with ThriveAtlas themes (ρ ranged from –0.21 to –0.39), indicating weak to moderate inverse correlations. The strongest relationships were observed for Overall Index (ρ = –0.39), Negative Life Experiences (ρ = –0.38), and , Provider Shortages (ρ = –0.38), with a notable correlation for Limited Wellness Practices (ρ = –0.32).

**Figure 2.**
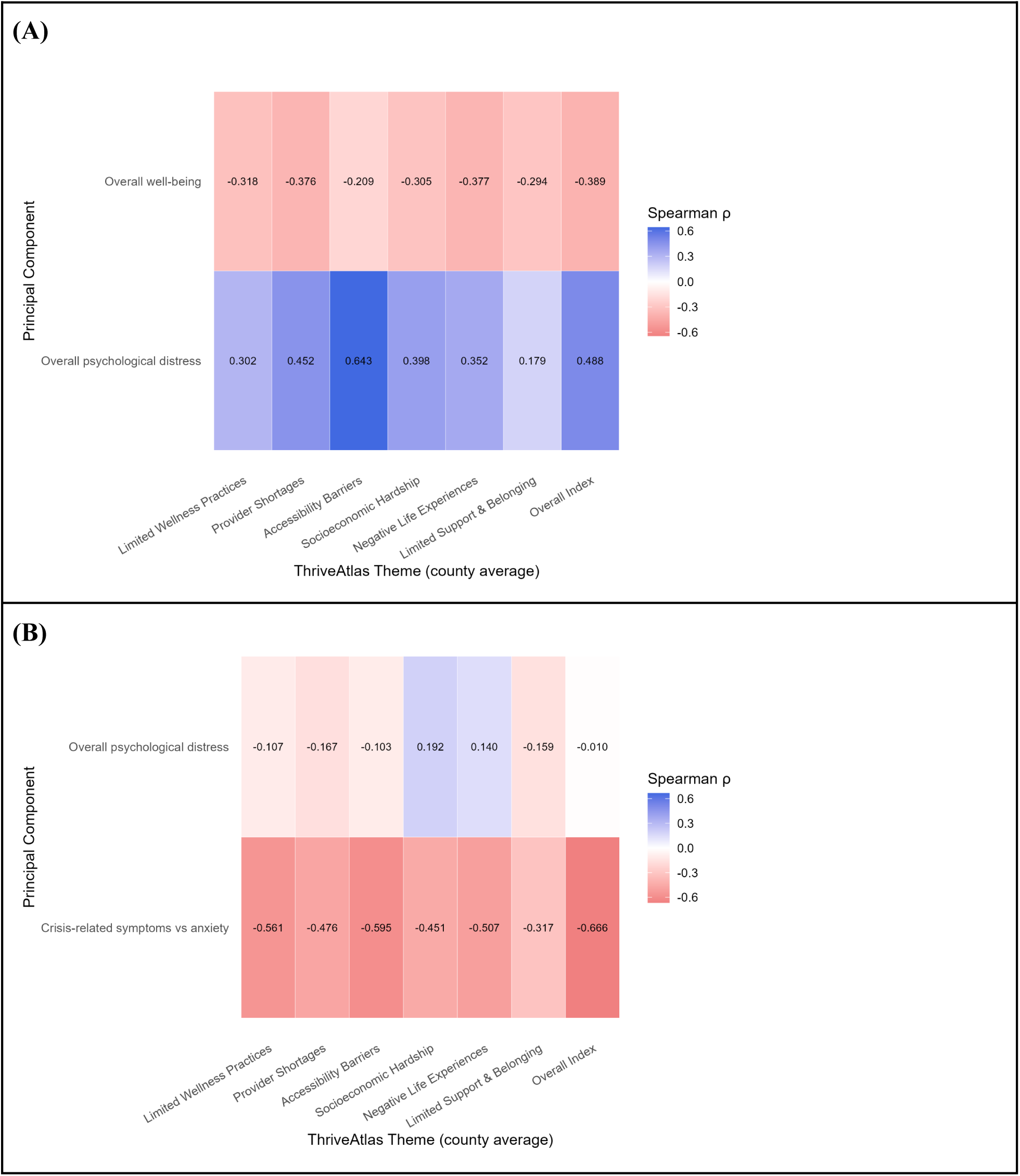
County-level Spearman correlations between ThriveAtlas Themes and principal components of youth mental health outcomes (A) California, Grade 9 (California Healthy Kids Survey, 2021–2023), (B) Washington, Grade 10 (Washington State Healthy Youth Survey, 2023). Principal components were derived separately for each state using state-specific youth mental health indicators. Component interpretations therefore differ across panels.

In Washington, overall distress (Component 1) exhibited weak and inconsistent associations with county-level ThriveAtlas themes (ρ ranged from –0.17 to 0.19). The largest positive correlations emerged for Socioeconomic Hardship (ρ = 0.19), suggesting slightly higher distress in counties with greater socioeconomic adversity. Notable negative correlations were observed for Provider Shortage (ρ =- 0.17) and Limited Support and Belonging (ρ = -0.16), indicating that counties with greater provider scarcity and lower levels of social support tended, unexpectedly, to report slightly lower levels of distress. Component 2, which differentiates crisis–related outcomes (e.g., suicide attempts, suicide plans, lack of adult support) from anxiety-related outcomes (e.g., anxiety, worry), showed consistently negative and substantially larger correlations across all ThriveAtlas themes, ranging from –0.32 to –0.67. Because crisis indicators load negatively and anxiety indicators load positively, negative correlations indicate that greater vulnerability was correlated with counties falling closer to the crisis end of the continuum. The strongest correlations for Component 2 were observed for the Overall Index (ρ = –0.67), Accessibility barriers (ρ = –0.60), Limited Wellness Practices (ρ = –0.56), Negative Life Experiences (ρ = –0.51), suggesting that higher vulnerability corresponds to a shift toward those experiencing crises rather than anxiety.

## Discussion

Our study highlights a clear and persistent gap in youth mental health and wellbeing measurement. Although numerous U.S. surveys and tools monitor youth mental health burden and related risk and protective factors, most function as surveillance tools and do not integrate geographically patterned drivers into an actionable measure of vulnerability. Among the limited tools that do consider these drivers, most capture only a narrow set of mental health indicators or associated drivers. Furthermore, many indices often emphasize general well-being domains (e.g., education, physical health, economic conditions), omitting determinants most relevant to youth mental health. Moreover, many of these tools offer information at course geographic scales (e.g., state), obscuring meaningful variations across neighborhoods and within communities. As a result, existing tools offer limited guidance for policymakers and stakeholders seeking to identify vulnerability, target resources, or identify actionable intervention levers. To address these gaps, we developed ThriveAtlas, a youth-centered index that integrates multiple drivers of youth mental health and well-being to support evidence-based, locally actionable decision-making.

ThriveAtlas directly addresses this gap by providing an index of relative vulnerability at the census tract-, zip code-, county-, and state-levels across the U.S. For analytic purposes, we examined county-level data and observed substantial variability nationwide, with elevated vulnerability appearing both in regional clusters and isolated counties, underscoring that the crisis is not confined to a single region and reflects diverse underlying drivers.^4,5^ These patterns highlight the need for targeted, place-based strategies in high-vulnerability clusters alongside capacity-building in counties where elevated need may otherwise be overlooked or obscured. By integrating multiple modifiable drivers of risk into the index’s subthemes, ThriveAtlas can help efficiently identify context-specific leverage points for prevention and intervention across communities.

Our validation analyses provide early support for the relevance and utility of ThriveAtlas, while also indicating that its observable relationship with youth mental health may vary by state. In California, associations followed expected patterns, with higher vulnerability corresponding to greater distress and lower well-being, supporting the construct validity of the index. In Washington, however, the pattern differed. Overall distress demonstrated weak and inconsistent correlations with ThriveAtlas themes, suggesting that ThriveAtlas captures only a subset of the factors associated with broad emotional distress relevant in this state. This divergence may reflect known limitations of self-reported mental health measures, which capture subjective experiences of distress and are shaped by reporting context, survey design, individual perception, and differential diagnosis and service contact, and therefore may not align closely with structural factors captured in ThriveAtlas.^53–55^ For example, counties with greater mental health provider shortages and limited social support tended to report slightly lower average distress, a counterintuitive finding likely reflecting underrecognition or underreporting, possibly due to rurality, stigma, or limited mental health literacy, rather than lower underlying need.^56–58^ By contrast, the crisis-related outcome aligned strongly with ThriveAtlas themes, suggesting that the index is particularly well aligned with identifying areas where youth mental health needs may be especially salient for crisis response.

Importantly, ThriveAtlas was developed to capture meaningful but inherently partial variation in youth mental health vulnerability. Youth mental health reflects a complex, dynamic system of interacting forces, many of which extend beyond the drivers included in this index due to conceptual scope and data availability.^14^ These include individual-level buffering and vulnerability factors (e.g., coping skills, temperament, genetics, substance use), interactions among risks (e.g., socioeconomic hardship moderated by family support and school climate), environmental determinants, and broader cultural, social, and psychological processes (e.g., social comparison, social media exposure, identify formation, hedonic adaptation).^17,59,60^ Accordingly, ThriveAtlas is designed to inform decision-making by identifying contexts in which multiple, empirically supported risk factors converge, rather than to predict mental health outcomes or estimate causal effects. ThriveAtlas should therefore be interpreted as a decision-relevant signal to support prioritization, prevention planning, and equity-oriented resource allocation, rather than outcome prediction.

This study has several limitations. First, the literature review did not follow systematic review procedures, and single-reviewer screening may have resulted in missed literature. Second, the index does not include all factors relevant to youth mental health; future iterations of ThriveAtlas may incorporate additional drivers as data availability improves. Third, validation analyses were limited to Washington and California, which were selected due to data availability and alignment with our study’s objectives. While this enabled a pragmatic assessment of alignment across distinct data contexts, data limitations in other states constrained broader multi-state validation, which remains an important direction for future work. Furthermore, although ThriveAtlas can be used down to the census tract level, we used county-level data for this analysis. County-level outcome data may mask substantial within-county variation in youth mental health outcomes. We were unable to find publicly accessible datasets that report youth mental health outcomes at finer geographic resolutions (e.g., ZIP code or census tract) that reliably provide relevant estimates. Many candidate datasets suppress results at sub-county levels, and in some cases even county levels, due to small sample sizes and privacy protections. Finally, ThriveAtlas captures a snapshot of structural conditions and does not account for temporal dynamics or resilience-promoting factors that may buffer youth mental health risk over time.

## Conclusion

Existing indices are often single-dimensional or insufficiently granular to inform targeted intervention and policy prioritization for youth mental health. ThriveAtlas addresses these gaps by offering a multidimensional, youth-centered, and geographically precise measure of vulnerability that reflects the contexts in which adolescents live and interact, with early validation supporting its utility for identifying communities facing elevated barriers to thriving. By translating complex, contextual determinants into interpretable and actionable metrics, ThriveAtlas offers stakeholders and youth-serving systems practical guidance for prioritizing investments, targeting prevention and early-intervention efforts, and advancing strategies responsive to local needs. With continued refinement as data availability improves, ThriveAtlas has the potential to inform policy, optimize resource allocation, and strengthen coordinated efforts to promote youth mental health and well-being.

## Acknowledgements

No acknowledgements.

