## Supplemental Files for "Measuring Youth Mental Health Vulnerability Across Communities: Development and Validation of ThriveAtlas"

**Supplemental Material**

### Supplemental Material Section 1 (S1)

### Small area estimation (SAE)

Many nationally representative surveys are not designed to produce reliable estimates for small geographic areas such as census tracts, ZIP Code Tabulation Areas (ZCTAs), or counties. To address this limitation, we use SAE methods to generate stable, local-level estimates by combining rich survey microdata with detailed population data from the U.S. Census.[1] Our approach leverages established statistical techniques to model individual-level outcomes and then re-weight those predictions to reflect the demographic composition of local communities. This allows us to produce consistent, comparable indicators of youth and young adult well-being across multiple geographic scales.

#### SAE using Behavioral Risk Factor Surveillance System (BRFSS)

We created eight indicators through a process of small-area estimation using techniques from the literature on Multilevel Regression and Poststratification.[2] We ingested microdata from the Behavioral Risk Factor Surveillance System (BRFSS) 2022 vintage dataset and created eight binary variables. To maintain relevance to youth outcomes, we limited the dataset to respondents aged 18 (the survey minimum) to 34. We then fit a random effects model to predict each variable, estimating random intercepts for respondents' state & census division, race/ethnicity, 5-year age group, education level (4 categories), with individual-level fixed effects for marital status, health insurance status, homeownership, and English-speaking ability.

We next generated individual predicted probabilities for each outcome using the American Community Survey 5-year Public Use Microdata Sample (PUMS), 2022 vintage, harmonized to match BRFSS features precisely and limited to individuals aged 18-24. Each PUMS individual is assigned to a Public Use Microdata Area (PUMA), a small area of about 100,000 residents; using PUMS as an inferential dataset allows us to account for small-area variations in influential covariates like educational achievement, marital status, and health insurance status, among others.

Using the PUMS inference dataset, we performed poststratification to census tract and Zip Code Tabulation Area (ZCTA) geographies using race/ethnicity (6 categories) X sex (2 categories) cells (12 cells in all) for each small area. Normally, age cells would also be used, but the narrow age band of our inference dataset precluded this. Once tract- and ZCTA-level estimates were generated, we performed population-weighted aggregation to arrive at county- and state-level estimates.

### SAE using National Survey of Children's Health (NSCH)

For the National Survey of Children's Health we created 4 indicators using NSCH microdata via a similar strategy to the BRFSS but with several key modifications. First, the NSCH is considerably smaller than the BRFSS microdata, so to fit a stable model we combined years 2019-2022 microdata. Second, the random and fixed effects are different for this survey. Child random effects: age group (2 categories), race/ethnicity (6 categories), sex (2 categories). Household fixed effects: language spoken, SNAP status, veteran status, highest education level achieved in the household, number of children under 5, whether a female or male (separately) adult was typically present, and homeownership.

Third, PUMS data only records adult (household) responses; we therefore limit inference to PUMS individuals listing a youth aged 10-17 and use harmonized household, parent, and child features as links between the NSCH source data and PUMS inference data.

### Supplemental Material Section 2 (S2)
#### Surgo Health Provider Database

Surgo Health's Provider Database is a US-focused repository tracking 6.1 million healthcare providers across 1.7 million healthcare organizations. For each provider, it aggregates their medical and pharmacy claims over the past 12 months, creating a provider care profile. These then create indicators around the density and variety of offered care.

### Supplemental Material Section 3 (S3)
#### Correlation and Principal Component Analysis for California and Washington State Validation

The supplemental figures and table below present county-level relationships among adolescent mental health outcomes in California and Washington using two complementary analytic approaches: correlation analysis and principal components analysis (PCA). First, Spearman rank correlations were used to examine how individual mental health outcomes co-vary across counties within each state. This nonparametric approach was selected to accommodate the non-normal distributions of the survey-derived prevalence measures. Correlation heatmaps summarize the extent to which different indicators, such as depressive feelings, anxiety symptoms, suicidality, and well-being, tend to increase or decrease together at the county level.

Second, PCA was used as a descriptive tool to summarize patterns of shared variation across multiple, correlated mental health outcomes within each state. We used the outcomes from the PCA, as the primary outcomes for our validation analysis. PCA reduces the dimensionality of the data by identifying latent components that capture common underlying constructs (e.g., overall emotional distress or well-being), while retaining most of the original variance. Scree plots are used to guide component selection, and factor loadings indicate the strength and direction with which each outcome contributes to a given component. Together, these analyses provide a structured view of how adolescent mental health outcomes cluster across counties and how distinct dimensions of distress, well-being, and crisis-related experiences emerge within each state. We have presented the results by state.

**Figure S1. California correlations between ThriveAtlas and county-level Grade 9 Mental Health outcomes (California Healthy Kids Survey 2021- 2023)**

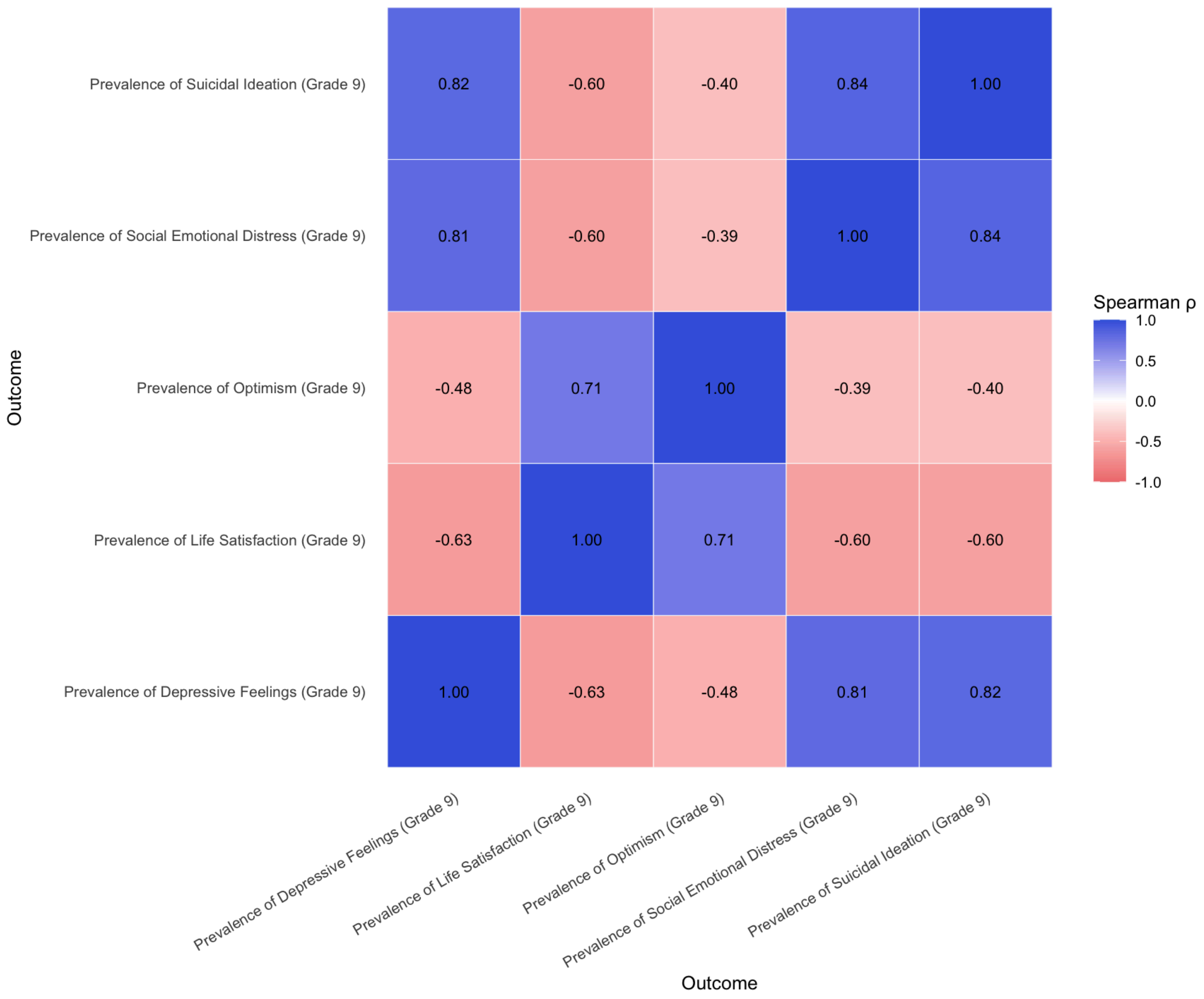

**Figure S2. Screeplot of Explained Variance by Principal Components**

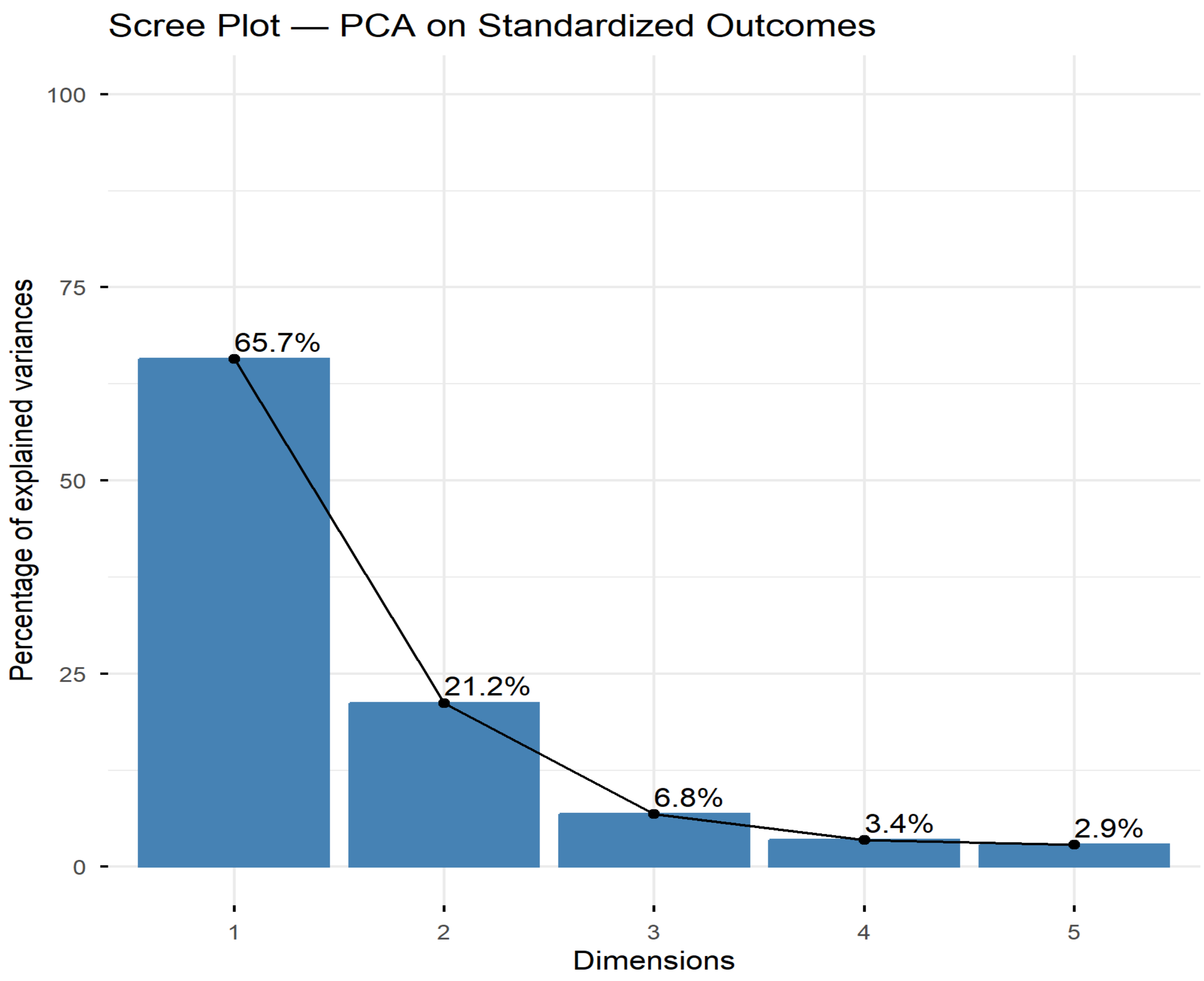

**Table S1. Factor Loadings for Component 1 and 2 in California**

| Outcome (County-level Prevalence) | Loading |
|---|---|
| **Component 1: Overall Distress (Grade 9)** | |
| Suicidal Ideation | 0.50 |
| Depressive Feelings | 0.50 |
| Social Emotional Distress | 0.47 |
| Life Satisfaction | -0.42 |
| Optimism | -0.31 |
| **Component 2: Overall Well-being (Grade 9)** | |
| Optimism | 0.74 |
| Life Satisfaction | 0.44 |
| Social Emotional Distress | 0.40 |
| Suicidal Ideation | 0.25 |
| Depressive Feelings | 0.22 |

**Figure S3. Washington correlations between ThriveAtlas and county-level Grade 10 Mental Health outcomes (Washington State Health Survey 2023)**

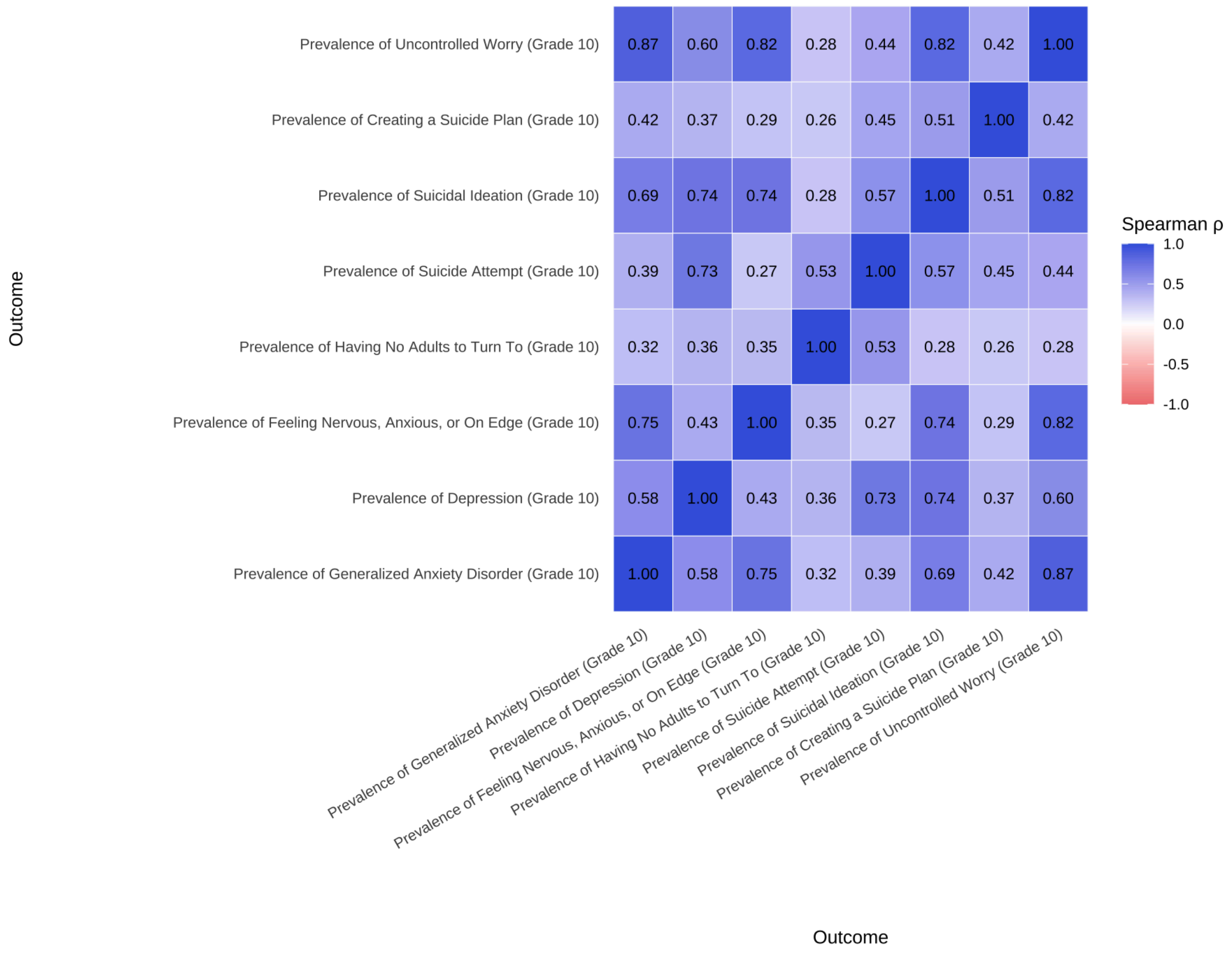

**Figure S4. Screeplot of Explained Variance by Principal Components**

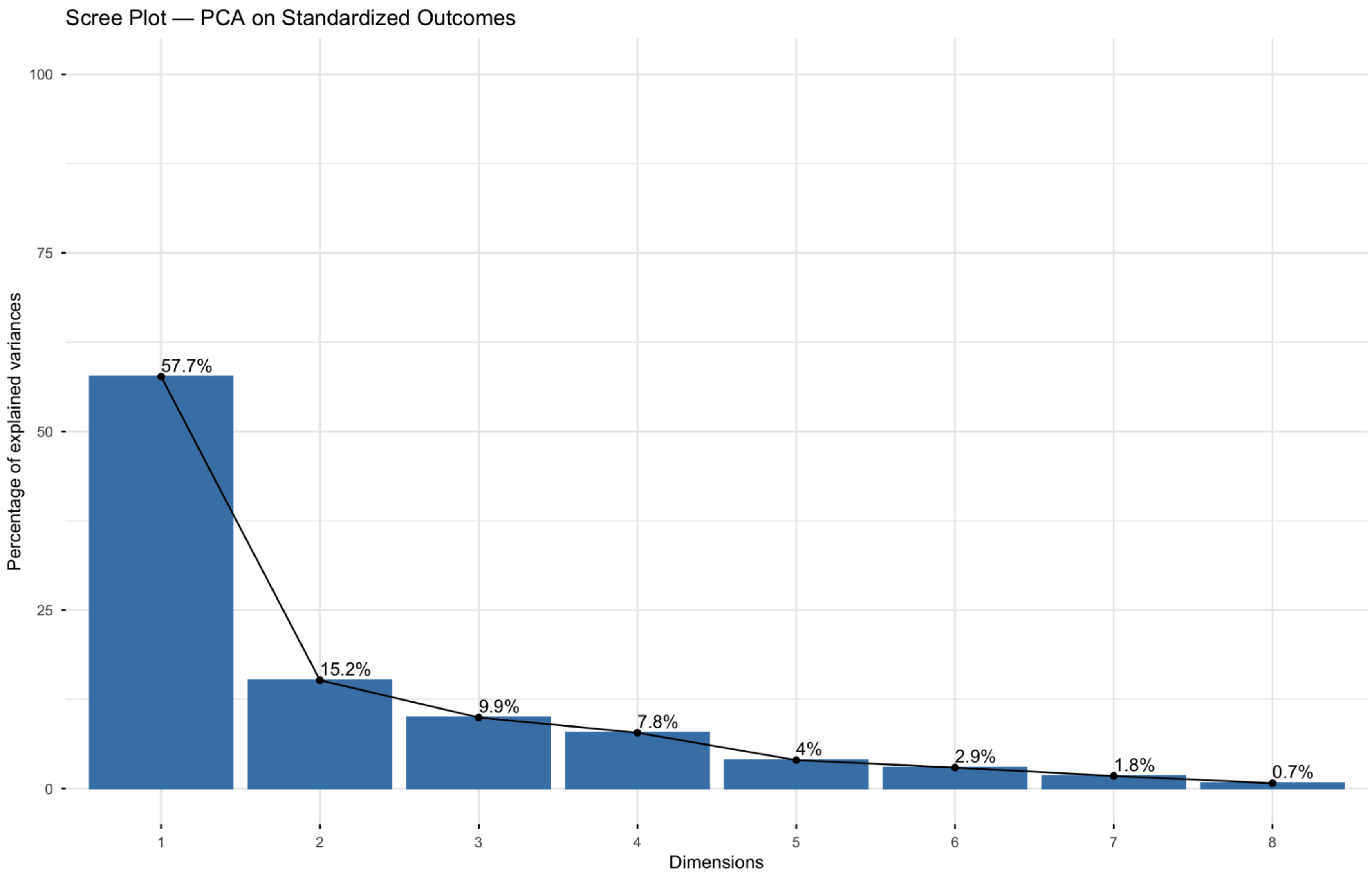

**Table S2. Factor Loadings for Component 1 and 2 in Washington**

| Outcome (County-level Prevalence) | Loading |
|---|---|
| **Component 1: Overall Distress (Grade 10)** | |
| Suicidal Ideation | -0.42 |
| Uncontrolled Worrying | -0.41 |
| Generalized Anxiety | -0.40 |
| Depressive Feelings | -0.37 |
| Nervous, Anxious, or on Edge | -0.35 |
| Suicide Attempt | -0.33 |
| Suicide Plan | -0.37 |
| No Adults to Report Feelings of Depression | -0.25 |
| **Component 2: Behavioral Crisis vs. Emotional Distress (Grade 10)** | |
| Suicide Attempt | -0.53 |
| No Adults to Report Feelings of Depression | -0.51 |
| Nervous, Anxious, or on Edge | 0.43 |
| Uncontrolled Worrying | 0.33 |
| Generalized Anxiety | 0.27 |
| Depressive Feelings | -0.22 |
| Suicide Plan | -0.17 |

| **Outcome (County-level Prevalence)** | **Loading** |
| --- | --- |
| Suicidal Ideation | 0.09 |
